# Bayesian, Universal COVID Testing

**DOI:** 10.1101/2021.04.23.21255984

**Authors:** Christian T. Meyer, Joel M. Kralj

## Abstract

During the SARS-COV2 pandemic, there has been a persistent call for universal testing to better inform policy decisions. However, a little considered aspect of this call is the relationship between a test’s accuracy and the tested demographic. What are the implications of frequent, universal testing in otherwise asymptomatic demographics? By applying Bayesian statistics, it becomes clear that as the odds of having COVID decreases, there is a non-linear increase in the odds that each positive test is, in fact, a false positive. This phenomenon has precedence in the historical narrative surrounding universal mammogram screening which is no longer recommended due to the unacceptably high rate of false positives. The solution to combat the inflation of false positives is also suggested by Bayesian statistics: intelligently integrating multiple COVID diagnostic tests and symptoms via Bayes’ Theorem, an approach conceptually similar to pre-screening for mammograms. This extra information is readily available (*e*.*g*. olfactory function and fever) and will minimize the economic and emotional costs incurred by false positives while simultaneously improving the information available for policy-makers. In summary, along with the push for universal testing should be an equally rigorous approach to interpreting the test results.

## Potential Risks of Universal Testing

Despite the extensive discussion surrounding various testing modalities for detecting the SARS-COV2 virus, there is comparatively little discourse on mitigating the impact of false positives [1] which exist for every test, regardless of the quality. The emotional and economic cost of false positives for universal testing has already been extensively discussed for universal mammogram screening, where it was realized that pre-screening significantly improved the diagnostic value of the test while minimizing the emotional burden of false positives [2]. In the case of mammograms, even though the nominal false-positive rate for a mammogram is less than 3% [3], universal screening resulted in approximately 38,057 false-positive mammograms out of 405,191 total tests [4], nearly four times the expected number of false positives. Just as for mammograms, the economic and human cost of every false positive in COVID tests is significant [1]. Children, extended families, and teachers are quarantined. Colleagues are forced to take unpaid time off and businesses are left with minimal staff. Entire buildings shut down for cleaning. Beyond the economic burden, a false positive promotes risky behavior by actors assuming immunity, thereby undermining the herd immunity [5].

The mammogram narrative suggests false positives are particularly problematic when historically diagnostic tests are being used for surveillance [6]. This is because the accuracy of the test decreases proportionally to the prior odds of the tested individual having the disease. This can be intuited from the following thought experiment. If you apply a test with a false positive rate of 1%, to a population of 100 people who do not have COVID, by definition you would have 1 false-positive and 0 false negatives. That means that 100% of the positives are false. Therefore, as you decrease the probability that your tested population has COVID by expanding the testing pool, you increase the proportion of positives which are false positives. Many doctors intuit this trade-off and understand that ordering a test is predicated on having a rationale for doing so (*i*.*e*. the patient is symptomatic).

## Bayesian Statistics Applied to COVID testing

This relationship between test performance and tested demographic can be formalized using Bayesian statistics, a branch of statistics dealing probability of an outcome given prior odds of an event. Formally, the probability of a false positive (*P* (*false*+))is equal to

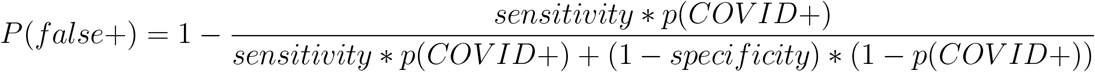

where specificity is the rate of true negatives (*p*(*test* −|*COVID* −)), sensitivity is the rate of true positives (*p*(*test* +| *COV ID*+)), and *p*(*COVID*+) is the probability of having COVID before the test (See Supplement for derivation). It is important to note that generally the sensitivity and specificity are not equal for any given test.

Examining the relationship between the percentage of positive tests which are false positives and the prior odds of having COVID reveals that as the prior odds of having COVID decreases, the proportion of false positives increases (Figure 1A) matching our expectation from the thought experiment. The proportion of false positives should not be confused with the *rate* of positives which decreases as the prior odds decrease. Increasing the specificity and sensitivity are not the solution as this trend is true of all tests to varying degrees (Figure 1A, B). This is not merely a curious phenomenon at some extreme, but plays an escalating role as the testing pool is continuously expanded. For example, taking the specifications of the gold standard PCR-based test which has 70-90% sensitivity and a remarkable 99% specificity, applied to a population with 1% prior odds of having COVID (estimated based on active caseload in a given area), false positives account for 52% of all positive tests.

**Figure 1:**
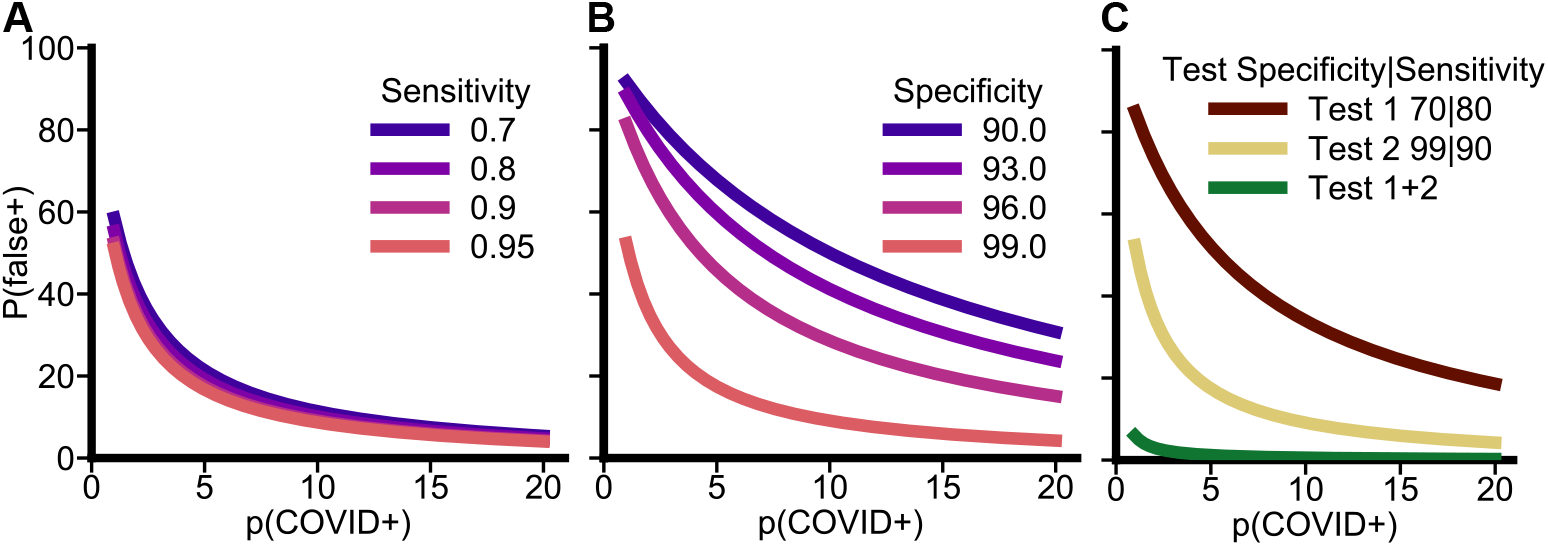
Diagnostic test performance depends on the prior odds of infection in the tested demographic. A,B) The probability of a false positive (*P* (*false*+)) depends on the prior odds of having COVID (*p*(*COV ID*+)) before the test. These odds necessarily decrease as the tested pool is expanded to more asymptomatic people. This is true even as sensitivity (A) or specificity (B) of a test increases, though test specificity is more important for false positives than sensitivity. C) Combining two tests (dark red and gold) of different sensitivity and specificity using a Bayesian approach, reduces the proportion of false positives (green). This is because the prior (*p*(*COV ID*+)) of the second test is updated with the posterior odds from the first test.

## Intelligently integrating multiple tests reduces the proportion of false positives

What is a solution to this tension? The shape of the curves in Figure 1A suggests even incremental increases in the prior odds dramatically reduce the probability of a false positive. When a test is being applied universally, the only approach to increasing the prior odds is to chain multiple tests together. This is because the outcome of each test becomes the prior odds for the subsequent test. Bayesian statistics, therefore, provides a simple solution to integrate multiple tests and reduce the economic burden of false positives.

For example, pairing an olfactory performance test [7] with PCR test has the following impact. If the olfactory test has specificity and sensitivity of 96% and 70% respectively [7], administered in a population with 1% prior odds of COVID+ results in false positives comprising 85% of the positives. If however, the positives are subsequently tested using the PCR test, the increase in prior odds from 1% to 18% based on the olfactory test decreases the proportion of false positives from 52% to 6% in the PCR test (Figure 1C, gold to green line). Because the order of the test does not matter, chaining together tests in order of the speed of turn around can improve the quality of each subsequent test while reducing the quarantine time for false positives thereby mitigating their economic and emotional impacts. Additionally, the posterior odds measured by each test decay back to the basal *p*(*COVID*+) level, as defined by the active caseload, over time. Therefore, reducing the spacing between subsequent tests is critical to maximizing information transfer between tests.

One could easily imagine extending this framework to include multiple COVID symptoms (*e*.*g*. fever, shortness of breath, etc.) in such a way that no information is wasted in identifying true positive cases. Indeed this is being done using Bayesian networks [8], though dealing with temporally sequenced and sparse information is critical for continuously updating diagnosis in real-time in a large population.

In summary, holistic assessment of the patient, rather than depending on a single test will always necessary in medicine, whether for breast cancer screening or COVID testing. That the quality of routine PCR tests could be greatly improved by simply appending 2-3 readily measurable symptoms to each test should be sufficient motivation to do so. Intelligent universal testing increases the value of the test while decreasing the economic and human costs of every false positive.

## Supplementary Information

### Bayes Derivation

Bayes’ equation for a the probability of being COVID+ given a positive test (*p*(*COVID* +| *test*+)), also called the positive predictive value, is

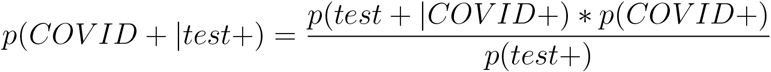

where *p*(*test* +|*COVID*+) is the probability of a positive test if COVID+ (commonly called a test’s **sensitivity**), *p*(*COVID*+) is the prior probability of having COVID before the test, and *p*(*test*+) is the probability of a positive test. The probability of a positive test is equal to

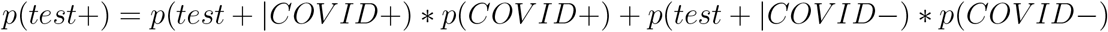

where *p*(*test* +|*COVID*−) is the probability of a positive test if one does not have COVID, commonly called the false positive rate. The **specificity** of a test is one minus the false positive rate (1 −*p*(*test* +|*COVID*−)). So rewriting the first equation to calculate the probability of false positive (*P* (*false*+) or *p*(*COVID-test*+) which is equal to 1 *-p*(*COVID* +|*test*+)) in terms of a test’s specificity and sensitivity results in the following equation

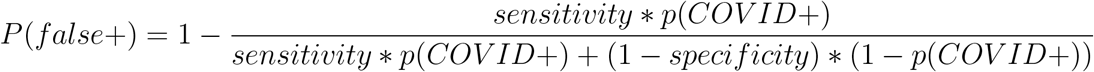

## Data Availability

Code required to recreate the analysis is available on request from lead author.

## Data Availability

Code required to recreate the analysis is available on request from lead author.

